# Vaccine effectiveness of CanSino (Adv5-nCoV) COVID-19 vaccine among childcare workers – Mexico, March–December 2021

**DOI:** 10.1101/2022.04.14.22273413

**Authors:** Vesta L. Richardson, Martín Alejandro Camacho Franco, Aurora Bautista Márquez, Libny Martínez Valdez, Luis Enrique Castro Ceronio, Vicente Cruz Cruz, Radhika Gharpure, Kathryn E. Lafond, Tat S. Yau, Eduardo Azziz-Baumgartner, Mauricio Hernández Ávila

## Abstract

**Background:** Beginning in March 2021, Mexico vaccinated childcare workers with a single-dose CanSino Biologics (Adv5-nCoV) COVID-19 vaccine. Although CanSino is currently approved for use in 10 Latin American, Asian, and European countries, little information is available about its vaccine effectiveness (VE).

**Methods:** We evaluated CanSino VE within a childcare worker cohort that included 1,408 childcare facilities. Participants were followed during March–December 2021 and tested through SARS-CoV-2 RT-PCR or rapid antigen test if they developed any symptom compatible with COVID-19. Vaccination status was obtained through worker registries. VE was calculated as 100% × (1−hazard ratio for SARS-CoV-2 infection in fully vaccinated vs. unvaccinated participants), using an Andersen-Gill model adjusted for age, sex, state, and local viral circulation.

**Results:** The cohort included 43,925 persons who were mostly (96%) female with a median age of 32 years; 37,646 (86%) were vaccinated with CanSino. During March–December 2021, 2,250 (5%) participants had laboratory-confirmed COVID-19, of whom 25 were hospitalized and 6 died. Adjusted VE was 20% (95% CI = 10–29%) against illness, 76% (42–90%) against hospitalization, and 94% (66–99%) against death. VE against illness declined from 48% (95% CI = 33–61) after 14–60 days following full vaccination to 20% (95% CI = 9–31) after 61–120 days.

**Conclusions:** CanSino vaccine was effective at preventing COVID-19 illness and highly effective at preventing hospitalization and death. It will be useful to further evaluate duration of protection and assess the value of booster doses to prevent COVID-19 and severe outcomes.

**Summary:** We evaluated CanSino (Adv5-nCoV) COVID-19 vaccine effectiveness during March–December 2021 using a childcare worker cohort that included 43,925 participants across Mexico. Vaccination decreased the risk of COVID-19 illness by 20%, hospitalization by 76%, and death by 94%.

## BACKGROUND

The CanSino Biologics and Beijing Institute of Biotechnology (CanSino) Ad5-nCoV COVID-19 vaccine is a single-dose adenovirus type 5 (Ad5)-vectored vaccine expressing the SARS-CoV-2 spike glycoprotein [1]. As of March 2022, the vaccine is approved for use in ten Latin American, Asian, and European countries (Argentina, Chile, China, Ecuador, Hungary, Indonesia, Malaysia, Mexico, Pakistan, and Republic of Moldova) and currently is under review for emergency use listing by the World Health Organization [2]. The CanSino vaccine is administered as a single intramuscular dose, is typically well-tolerated, and induces neutralizing antibodies against SARS-CoV-2 [3-5]. Limited data suggest that the CanSino vaccine is less immunogenic than mRNA vaccines but consistent with other viral vector vaccines [6, 7]. Additionally, phase 3 clinical trial results at 28 days after CanSino vaccination have demonstrated a vaccine efficacy of 58% [1]. Otherwise, little is published about CanSino’s real-world effectiveness in preventing illness, hospitalization, and death; two studies from Malaysia and Pakistan included patients vaccinated with CanSino in national estimates demonstrating vaccine impact [8, 9], but no product-specific vaccine effectiveness estimates have been published to date.

On February 9, 2021, Mexico’s Federal Commission for the Protection against Sanitary Risks authorized the CanSino vaccine for emergency use in persons aged ≥18 years to mitigate the effect of the COVID-19 pandemic [10]. Throughout the pandemic, middle income countries like Mexico struggled to secure timely supplies of well-studied vaccines, such as mRNA vaccine products, and often relied on relatively understudied products [11]. Mexico obtained CanSino vaccine because it is administered in one dose and can be stored at 2–8°C [12], which facilitates handling, storage, and distribution throughout Mexico’s Universal Vaccination Program network. Health authorities initiated the COVID-19 vaccination campaign on March 15, 2021, by offering voluntary, free-of-charge, CanSino vaccines to school workers, including childcare workers, for several days at different official vaccination sites. Health authorities hoped that Mexico’s history of early adoption of immunizations coupled with vaccine promotion would achieve high CanSino coverage and effectively protect essential workers, such as childcare workers and teachers, as they returned to in-person work.

Understanding real-world effectiveness is especially important in middle-income countries where some jurisdictions might struggle with cold-chain and logistic challenges and vaccine access issues [11]. Furthermore, most currently available COVID-19 vaccines were tailored to produce immunogenicity against wild-type SARS-CoV-2 rather than currently circulating variants. Understanding current effectiveness and duration of protection, particularly in the context of novel emerging variants is a global priority and can inform health authorities’ strategies for homologous or heterologous boosting [13]. We examined data from a large cohort of childcare workers in Mexico to quantify the real-world effectiveness of the CanSino vaccine, an understudied product, in preventing illness, hospitalization, and death associated with COVID-19.

## METHODS

### Study Design

We evaluated the effectiveness of CanSino vaccine in a cohort of 56,483 Instituto Mexicano del Seguro Social (IMSS)-affiliated workers in 1,408 childcare centers across all 32 states of Mexico. The cohort was established in July 2020 to monitor COVID-19 illnesses at childcare centers after these began to reopen that month. Childcare workers were in daily contact with children aged ≤ 4 years, an age group that can readily transmit SARS-CoV-2 [14] and is currently ineligible for vaccination. IMSS promoted the use of masks and face shields among staff, frequent handwashing, alcohol-based gels, separation between groups of children and teachers in their classroom and for outdoor activities, limited classroom occupancy, and adequate ventilation, to reduce the risk of SARS-CoV-2 transmission in childcare settings. IMSS also established a mandatory active surveillance system to rapidly identify and isolate suspected COVID-19 cases among staff and children. Staff at childcare centers were instructed to maintain registries of workers’ basic demographic information (e.g., age and sex) which they subsequently used to each day to record attendance, COVID-19 status, and self-reported vaccination status. For persons with COVID-19-associated deaths, IMSS staff verified vaccine administration reported in these worker registries with entries in the national COVID-19 vaccination registry.

Each day during the five-day work week, staff obtained temperatures from children and workers in each childcare center. Additionally, three times a day, staff assessed whether anyone had developed signs or symptoms compatible with COVID-19. Staff also called workers who did not present for duty to determine if they had developed signs or symptoms. Workers who developed symptoms during the weekend were instructed to report these to their supervisor. IMSS defined a suspected case of COVID-19 as development of acute (<10 days) cough, fever, difficulty breathing, or headache in addition to one of the following signs or symptoms: arthralgias, myalgias, sore throat, loss of taste or smell, chest pain, chills, rhinorrhea, excessive lacrimation, vomiting, abdominal pain, or diarrhea. Workers with suspected COVID-19 were instructed to immediately isolate and seek nasopharyngeal swabbing for either reverse transcription polymerase chain reaction (RT-PCR) or rapid antigen SARS-CoV-2 testing at the nearest IMSS family clinic. Workers were then instructed to bring or send an image of the laboratory results to their supervisors.

Persons with suspected COVID-19 who then had a SARS-CoV-2-positive laboratory result were subsequently reclassified as laboratory-confirmed COVID-19 cases in the surveillance database. If workers developed COVID-19, IMSS staff periodically called or texted them to follow illness progression and convalescence. If workers were too sick to report, IMSS staff called their next of kin to follow up on illness progression. IMSS staff systematically recorded participant or proxy reports of hospitalizations associated with the COVID-19 illness and, when feasible, verified pre-existing conditions and outcomes with available records. Additionally, IMSS staff verified deaths associated with COVID-19 through a review of the Sistema de Notificación en Línea para la Vigilancia Epidemiológica (SINOLAVE) national surveillance system and death certificates. For patients who were hospitalized or who died, IMSS staff also gathered information about underlying health conditions including physician-diagnosed diabetes, high blood pressure, cardiovascular disease, obesity, chronic kidney disease, chronic obstructive pulmonary disease, pregnancy, or cancer from SINOLAVE.

### Statistical Analysis

We restricted our vaccine effectiveness analyses to participants who contributed person-time during March 30, 2021, the date when CanSino vaccine had been available for ≥14 days to childcare workers at IMSS, to December 31, 2021, the latest available data for the cohort. We also restricted our analysis to persons aged ≥18 years, the age of eligibility to the CanSino vaccine. We excluded cohort participants who reported a laboratory-confirmed COVID-19 illness prior to March 30, 2021 (n=1,074), were vaccinated with COVID-19 vaccine products other than CanSino (n=11,415) and developed COVID-19 within 13 days after vaccination with CanSino (n=69), resulting in 43,925 included cohort participants. We then classified participants into fully vaccinated with CanSino (≥14 days after receipt of CanSino vaccine) or unvaccinated. As in other vaccine effectiveness analyses, we considered the 13 days between vaccination and full vaccination as excluded person-time [15].

We estimated vaccine effectiveness for three outcomes: laboratory-confirmed COVID-19 illness, COVID-19–associated hospitalization, and COVID-19–associated death. Participant characteristics were first compared by vaccination status using Chi-square tests to explore propensity to vaccination. We then calculated the rolling 7-day daily incidence of laboratory-confirmed COVID-19 by vaccination status. We assessed which SARS-CoV-2 variant represented >50% of SARS-CoV-2 sequences during the study period using data submitted from Mexico to the Global Initiative on Sharing Avian Influenza Data (GISAID) [16]. We classified June 29–December 19 as B.1.617.2 (Delta) variant predominance based on GISAID data.

Hazard ratios and 95% confidence intervals for outcomes in fully vaccinated participants, as compared with unvaccinated participants, were estimated with the Andersen-Gill extension of the Cox proportional hazards model, which accounted for time-varying vaccination status (i.e., persons could contribute both unvaccinated and fully vaccinated person time). Unadjusted vaccine effectiveness was calculated with the following formula: 100% × (1−hazard ratio). An adjusted vaccine effectiveness model included a priori characteristics that could confound the association between vaccination and outcomes, namely age, sex, and the state in which the childcare center was located, in addition to local viral circulation, which was the weekly percentage positive of SARS-CoV-2 tests performed in the state, obtained via the Mexico Dirección General de Epidemiología public COVID-19 data dashboard [17]. We stratified vaccine effectiveness by days since full vaccination, and also calculated vaccine effectiveness prior to and during Delta variant predominance in Mexico. We did not calculate vaccine effectiveness post-Delta predominance due to limited follow-up time available (2 weeks). As a sensitivity analysis, we stratified vaccine effectiveness estimates by type of laboratory test used for confirmation (rapid antigen test vs. RT-PCR). All analyses were conducted with SAS software, version 9.4 (SAS Institute).

This IMSS-funded vaccine evaluation occurred within the context of emergency response and used anonymized workplace surveillance data. IMSS waived ethical approval for this work as the evaluation was a non-research, public health surveillance activity and both data collection and illness tracking were a requirement for working in these childcare centers during the pandemic.

## RESULTS

Cohort participants were primarily female (96%) and aged 18–49 years (90%) with a median age of 32 years (interquartile range [IQR] = 26–41) (Table 1). Among all 43,925 cohort participants, 37,646 (86%) were vaccinated with CanSino. We observed differences in age and geographic site, both in frequency of laboratory-confirmed illness and vaccination status, and also observed differences in sex by vaccination status (**Table 1** and **Supplementary Table 1**). Among persons who were hospitalized, 18 (72%) reported at least one underlying medical condition, including all 6 (100%) patients who died (**Supplementary Table 2**).

**Table 1:** Characteristics of participants by development of laboratory-confirmed COVID-19 and by vaccination status -- Mexico, 2021

| Characteristic | Total | % | Laboratory-confirmed COVID-19<sup>1</sup> |  |  |  | p-value | Vaccination status |  |  |  | p-value |
| --- | --- | --- | --- | --- | --- | --- | --- | --- | --- | --- | --- | --- |
|  |  |  | Yes | % | No | % |  | Vaccinated with CanSino<sup>2</sup> | % | Invaccinated | % |  |
| **Total** | 43925 | 100% | 2250 | 100% | 41675 | 100% | -- | 37646 | 100% | 6279 | 100% | -- |
| **Sex** |  |  |  |  |  |  |  |  |  |  |  |  |
| Female | 42056 | 96% | 2142 | 95% | 39914 | 96% | 0.19 | 36107 | 96% | 5949 | 95% | <0.0001 |
| Male | 1869 | 4% | 108 | 5% | 1761 | 4% |  | 1539 | 4% | 330 | 5% |  |
| **Age group** |  |  |  |  |  |  |  |  |  |  |  |  |
| 18-49 | 39533 | 90% | 2089 | 93% | 37444 | 90% | <0.0001 | 33669 | 89% | 5864 | 93% | <0.0001 |
| 50+ | 4392 | 10% | 161 | 7% | 4231 | 10% |  | 3977 | 11% | 415 | 7% |  |
| **Region<sup>3</sup>** |  |  |  |  |  |  |  |  |  |  |  |  |
| Central | 10149 | 23% | 519 | 23% | 9630 | 23% | <0.0001 | 8859 | 24% | 1290 | 21% | <0.0001 |
| North | 13816 | 31% | 508 | 23% | 13308 | 32% |  | 11233 | 30% | 2583 | 41% |  |
| West | 14509 | 33% | 789 | 35% | 13720 | 33% |  | 12728 | 34% | 1781 | 28% |  |
| South | 5449 | 12% | 434 | 19% | 5015 | 12% |  | 4825 | 13% | 624 | 10% |  |

<sup>1</sup>Both SARS-CoV-2 RT-PCR and rapid antigen tests were used for laboratory confirmation.
<sup>2</sup>Includes persons vaccinated with CanSino at any time during cohort follow-up from March–December 2021.
<sup>3</sup>Regions defined as: Central (Guerrero, Estado de México, Morelos, Querétaro, Distrito Federal); North (Aguascalientes, Coahuila, Chihuahua Durango, Nuevo León, San Luis Potosí, Tamaulipas, Zacatecas); West (Baja California, Baja California Sur, Colima, Guanajuato, Jalisco, Michoacán, Nayarit, Sinaloa, Sonora); South (Campeche, Chiapas, Hidalgo, Oaxaca, Puebla, Quintana Roo, Tabasco, Tlaxcala, Veracruz, Yucatán). Infection and vaccination status by state are presented in **Supplementary Table 1**.

The majority of cohort participants were fully vaccinated during May–June 2021, and incidence of COVID-19 among both fully vaccinated and unvaccinated participants peaked in August 2021 (**Figure 1**). In total, 2,250/43,925 (5%) participants developed laboratory-confirmed COVID-19, including 1,855/37,646 (5%) fully vaccinated and 395/6,279 (6%) unvaccinated persons. Of these 2,250 laboratory-confirmed cases, 2104 (94%) were diagnosed by rapid antigen test and 146 (6%) by RT-PCR; participants diagnosed by rapid antigen test vs. RT-PCR were comparable in sex, age, and vaccination status, but differed by state (**Supplementary Table 3**). Among fully vaccinated persons, the median time from vaccination to symptom onset was 104 days (IQR = 79–129). Twenty-five (0.06%) participants were hospitalized, including 14/37,646 (0.04%) fully vaccinated and 11/6,279 (0.18%) unvaccinated persons, and six (0.01%) participants died, including two (0.01%) fully vaccinated and four (0.06%) unvaccinated persons.

**Figure 1:** Timing of vaccination and incidence of laboratory-confirmed COVID-19 in the study cohort -- Mexico, 2021. Fully vaccinated was defined as ≥14 days after receipt of vaccine administration. Both SARS-CoV-2 RT-PCR and rapid antigen tests were used for laboratory confirmation. Delta variant predominance was defined as the time period during which >50% of SARS-CoV-2 sequences submitted to the Global Initiative on Sharing Avian Influenza Data (GISAID) were characterized as Delta variant (accessed via the PAHO SARS-CoV-2 Variants Tracking in the Region of the Americas dashboard [16]). COVID-19 incidence is presented as the rolling 7-day average among unvaccinated and fully vaccinated cohort participants.

During the study period, unvaccinated persons contributed a total of 3,164,516 person-days and fully vaccinated persons contributed 8,188,809 person-days (**Table 2**). The unadjusted vaccine effectiveness was 14% (95% confidence interval [CI] = 3–23) against laboratory-confirmed COVID-19, 73% (95% CI = 36–88) against COVID-19-associated hospitalization, and 92% (95% CI = 55–99) against COVID-19-associated death. After adjusting for age, sex, state, and local viral circulation, vaccine effectiveness over the full cohort follow-up period was 20% (95% CI = 10–29) against laboratory-confirmed illness, 76% (95% CI = 42–90) against hospitalization, and 94% (95% CI = 66–99) against death. When evaluating only persons diagnosed via RT-PCR, adjusted estimates were comparable to overall adjusted estimates but had low precision, with a vaccine effectiveness of 16% (95% CI = -36–48%) against illness and 75% (95% CI = -21-95%) against hospitalization (**Supplementary Table 4**).

**Table 2:**
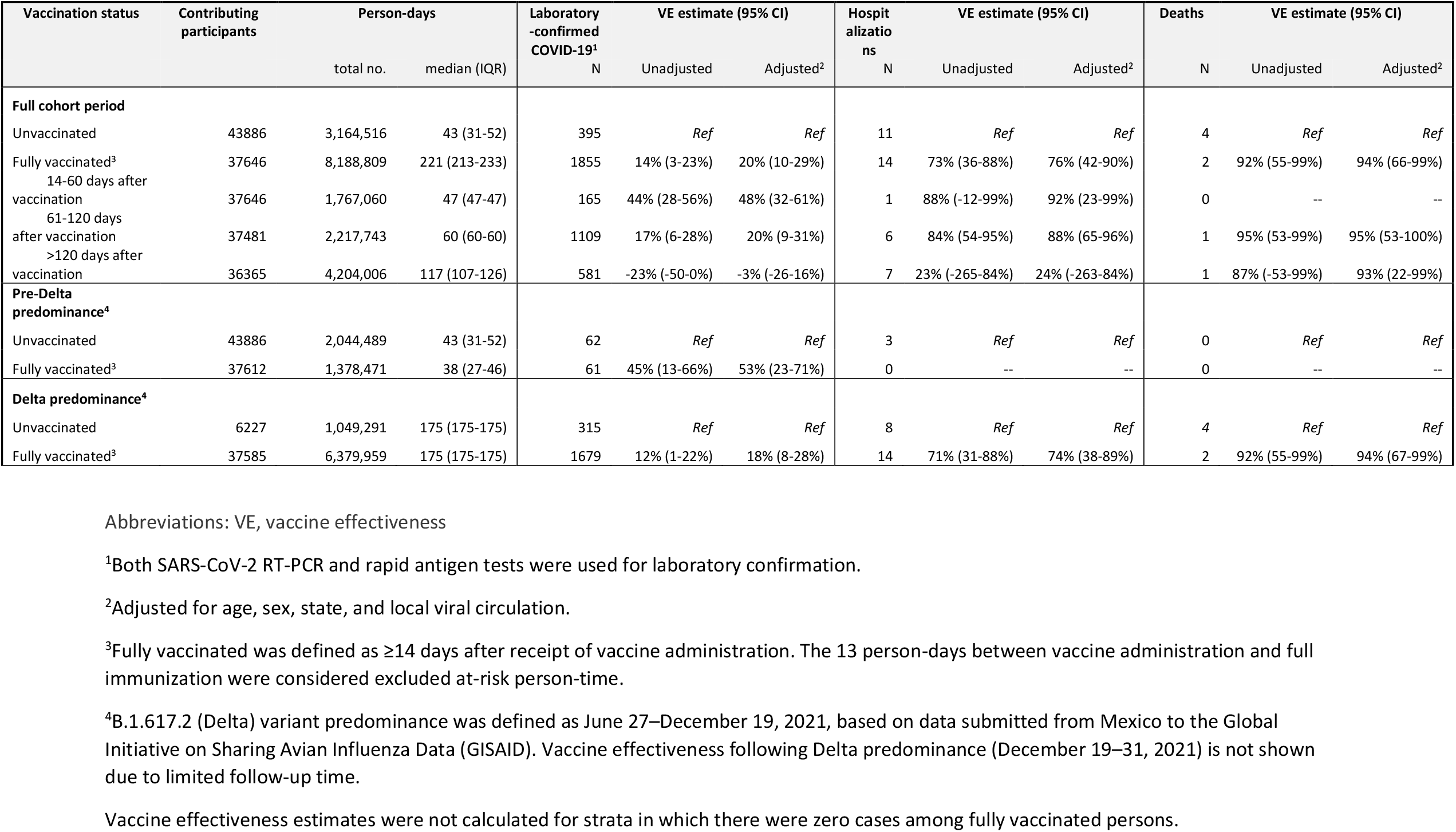
Effectiveness of CanSino vaccine in preventing laboratory-confirmed COVID-19, hospitalization, and death -- Mexico, 2021

Adjusted vaccine effectiveness against illness prior to Delta variant predominance (March 30– June 28, 2021) was 53% (95% CI = 23–71); this declined to 18% (95% CI = 8–28) during Delta predominance (**Table 2**). Additionally, vaccine effectiveness against illness decreased with longer time since vaccination; adjusted vaccine effectiveness declined from 48% (95% CI = 32–61) after 14–60 days following full vaccination to 20% (95% CI = 9–31) after 60–120 days following full vaccination. Vaccine did not confer significant protection against illness greater than 120 days following full vaccination (vaccine effectiveness = -3% [95% CI = -26–16%]). Vaccine effectiveness against hospitalization did not decline substantially in the first 120 days, with estimates at 92% (95% CI = 23–99) after 14–60 days following vaccination and 88% (95% CI = 65–96) after 60–120 days. However, protection against hospitalization was not significant after 120 days (vaccine effectiveness = 24% [95% CI = -263–84%]). No deaths were reported in fully vaccinated persons within 14–60 days of vaccination; however, vaccine effectiveness did not decline substantially from 61–120 days to greater than 120 days (95% and 93%, respectively).

## DISCUSSION

Our evaluation of the real-world effectiveness of the CanSino vaccine in Mexico suggests that most IMSS-affiliated childcare workers sought CanSino vaccines and had a 20% reduction in risk of COVID-19 illness, as well as a 76% reduction in risk of COVID-19-associated hospitalization and 94% reduction in risk of death. Peak incidence of cases in both vaccinated and unvaccinated cohort participants coincided with Delta variant emergence in Mexico, and vaccine effectiveness against illness was lower during Delta variant predominance compared with prior months. Nevertheless, vaccine effectiveness against hospitalization and death remained high during Delta predominance, consistent with prior reports of mild breakthrough infection with Delta among persons vaccinated with CanSino in Mexico [18]. However, Delta variant emergence also coincided with increasing time since vaccination, and as in similar prior studies, we could not distinguish between effects of both factors [19]. Our data indicated that CanSino vaccine was most effective early after administration and declined by 4 months after administration; vaccine did not appear to confer continued protection after 120 days following vaccination, though interpretation of vaccine effectiveness is limited by very wide confidence intervals. These findings were consistent with a recent systematic review demonstrating that other COVID-19 vaccines wane in their effectiveness by >20% during the first 1–6 months after administration [20].

Our early 14–60 day vaccine effectiveness estimate of 48% (95% CI = 32–60) was similar to the 28-day efficacy of single-dose CanSino vaccine against PCR-confirmed illness in the large multi-country clinical trial among persons aged ≥18 years which included study sites in Mexico (58% [95% CI = 40–70%] [1] and to preliminary unpublished 28-day vaccine effectiveness in Chile after two doses of CanSino (52% [95% CI = 49–55%]) [21]. Additionally, our overall vaccine effectiveness against COVID-19-associated hospitalization, at 76% (95% CI = 42–90), was also comparable to single-dose Janssen Ad26.COV2.S real-world vaccine effectiveness against COVID-19-associated hospitalization in the United States at 68% (95% CI = 50–79) [22].

It is likely that waning vaccine effectiveness over time is driven by antigenic drift as new variants evolve and develop new antigenic properties to evade existing antibodies, as well as waning of immune response over time [23-25]. Such findings have compelled many technical advisory groups to explore the value of homologous or heterologous boosting to better maintain the level of protection initially offered by mRNA vaccines [26, 27]. While effective, single-dose COVID-19 vaccination with products like CanSino tend to provide less protection than multi-dose schedules [28], and further analyses of this cohort of childcare workers might be useful in assessing the utility and optimal timing of homologous or heterologous boosting against SARS-CoV-2, as has been done with other single-dose COVID-19 vaccines [29-31].

### Strengths and limitations

Our evaluation demonstrated noteworthy strengths of IMSS’s worker surveillance and vaccine rollout in Mexico. We followed nearly 44,000 childcare workers during 2021 which allowed us to prospectively monitor COVID-19 illness development and ascertain subsequent hospitalization and death. High vaccine coverage (86%) among this cohort was facilitated by Mexico’s vaccination rollout through an existing universal vaccination program; assessments of previous pandemics demonstrate that countries that have existing immunization programs are more likely to rapidly benefit from pandemic vaccines than countries without such programs [32].

However, our study also had important limitations. Cohort members were predominantly female (96%) with a median age of 32 years, and findings may not be generalizable to other populations. Sparse outcomes, particularly hospitalizations and deaths, reduced the precision of vaccine effectiveness estimates. Additionally, a large proportion of workers with symptoms compatible with COVID-19 were tested through rapid antigen tests rather than through the more sensitive RT-PCR assays [33]; however, results of our sensitivity analysis indicated that estimates were comparable for overall and RT-PCR-only test results. Key data about SARS-CoV-2 infection and COVID-19 illness (including presence of symptoms, laboratory result, and vaccination status) were derived in many instances through self-report, although these were verified by official sources whenever possible. Finally, data on underlying medical conditions were only available for persons who were hospitalized or who died and could not be evaluated as a covariate in vaccine effectiveness models. Individuals with underlying conditions may have sought vaccination earlier than those without conditions, potentially causing differential risk from waning protection during the Delta wave in this population. As this was an observational study, unmeasured and residual confounding might have also been present.

## CONCLUSION

Our evaluation suggests that CanSino vaccine reduced the risk of COVID-19 illness among childcare workers by 20% and reduced risk of COVID-19-associated hospitalization and death by 76% and 94%, respectively. Like most COVID-19 vaccine products, vaccine effectiveness waned substantially during the first 4 months after administration, suggesting the potential value of homologous or heterologous booster doses. Additional vaccine effectiveness evaluations are warranted following Omicron SARS-CoV-2 variant predominance throughout Mexico and possible booster doses received by cohort participants.

## Data Availability

Data produced in the present study are available upon reasonable request to the authors.

## ACKNOWLEDGMENTS

We would like to thank the childcare workers and supervisors who participated in the COVID-19 surveillance and their families, who made this evaluation possible. We would also like to thank Daniel (Young) Yoo for his assistance with mathematical modeling and Drs. Alicia Fry and Michael Jhung for their expert review. The findings and conclusions in this report are those of the authors and do not necessarily represent the official position of the Centers for Disease Control and Prevention.

## Funding

This evaluation has been funded through the IMSS, Government of Mexico.

## Conflict of interest

None of the coauthors have conflicts of interest to declare.

## Notes

### Competing Interest Statement

The authors have declared no competing interest.

### Author Declarations

The Instituto Mexicano del Seguro Social waived ethical approval for this work as the evaluation was a non-research, public health surveillance activity and both data collection and illness tracking were a requirement for working in these childcare centers during the pandemic.

