## Supplementary for "Vaccine effectiveness of CanSino (Adv5-nCoV) COVID-19 vaccine among childcare workers – Mexico, March–December 2021"

**Supplementary Table 1: Participants with laboratory-confirmed COVID-19 and by vaccination status, by state -- Mexico, 2021**

| State | Total | % | Laboratory-confirmed COVID-19[1] | | | | | Vaccination status | | | | |
|---|---|---|---|---|---|---|---|---|---|---|---|---|
| | | | Yes | % | No | % | p-value | Vaccinated with CanSino[2] | % | Unvaccinated | % | p-value |
| **Total** | 43925 | 100% | 2250 | 100% | 41675 | 100% | <0.0001 | 37646 | 100% | 6279 | 100% | <0.0001 |
| Distrito Federal | 4635 | 11% | 343 | 15% | 4292 | 10% | | 4099 | 11% | 536 | 9% | |
| Aguascalientes | 860 | 2% | 31 | 1% | 829 | 2% | | 801 | 2% | 59 | 1% | |
| Baja California | 1789 | 4% | 56 | 2% | 1733 | 4% | | 1437 | 4% | 352 | 6% | |
| Baja California Sur | 434 | 1% | 59 | 3% | 375 | 1% | | 376 | 1% | 58 | 1% | |
| Campeche | 25 | 0% | 5 | 0% | 20 | 0% | | 0 | 0% | 25 | 0% | |
| Chiapas | 438 | 1% | 26 | 1% | 412 | 1% | | 362 | 1% | 76 | 1% | |
| Chihuahua | 3096 | 7% | 46 | 2% | 3050 | 7% | | 2305 | 6% | 791 | 13% | |
| Coahuila | 2043 | 5% | 72 | 3% | 1971 | 5% | | 1559 | 4% | 484 | 8% | |
| Colima | 586 | 1% | 39 | 2% | 547 | 1% | | 502 | 1% | 84 | 1% | |
| Durango | 582 | 1% | 24 | 1% | 558 | 1% | | 492 | 1% | 90 | 1% | |
| Estado De México | 2760 | 6% | 74 | 3% | 2686 | 6% | | 2366 | 6% | 394 | 6% | |
| Guanajuato | 2561 | 6% | 97 | 4% | 2464 | 6% | | 2405 | 6% | 156 | 2% | |
| Guerrero | 497 | 1% | 25 | 1% | 472 | 1% | | 483 | 1% | 14 | 0% | |
| Hidalgo | 423 | 1% | 29 | 1% | 394 | 1% | | 395 | 1% | 28 | 0% | |
| Jalisco | 2996 | 7% | 112 | 5% | 2884 | 7% | | 2469 | 7% | 527 | 8% | |
| Michoacán | 1477 | 3% | 109 | 5% | 1368 | 3% | | 1310 | 3% | 167 | 3% | |
| Morelos | 1009 | 2% | 35 | 2% | 974 | 2% | | 877 | 2% | 132 | 2% | |
| Nayarit | 610 | 1% | 83 | 4% | 527 | 1% | | 545 | 1% | 65 | 1% | |
| Nuevo León | 2884 | 7% | 150 | 7% | 2734 | 7% | | 2168 | 6% | 716 | 11% | |
| Oaxaca | 319 | 1% | 35 | 2% | 284 | 1% | | 289 | 1% | 30 | 0% | |
| Puebla | 749 | 2% | 31 | 1% | 718 | 2% | | 694 | 2% | 55 | 1% | |
| Querétaro | 1248 | 3% | 42 | 2% | 1206 | 3% | | 1034 | 3% | 214 | 3% | |
| Quintana Roo | 675 | 2% | 47 | 2% | 628 | 2% | | 535 | 1% | 140 | 2% | |
| San Luis Potosí | 1209 | 3% | 50 | 2% | 1159 | 3% | | 1076 | 3% | 133 | 2% | |
| Sinaloa | 1896 | 4% | 137 | 6% | 1759 | 4% | | 1666 | 4% | 230 | 4% | |
| Sonora | 2160 | 5% | 97 | 4% | 2063 | 5% | | 2018 | 5% | 142 | 2% | |
| Tabasco | 179 | 0% | 22 | 1% | 157 | 0% | | 154 | 0% | 25 | 0% | |
| Tamaulipas | 2396 | 5% | 108 | 5% | 2288 | 5% | | 2117 | 6% | 279 | 4% | |
| Tlaxcala | 153 | 0% | 4 | 0% | 149 | 0% | | 150 | 0% | 3 | 0% | |
| Veracruz | 1488 | 3% | 143 | 6% | 1345 | 3% | | 1340 | 4% | 148 | 2% | |
| Yucatán | 1000 | 2% | 92 | 4% | 908 | 2% | | 906 | 2% | 94 | 1% | |
| Zacatecas | 746 | 2% | 27 | 1% | 719 | 2% | | 715 | 2% | 31 | 0% | |

[1]Both SARS-CoV-2 RT-PCR and rapid antigen tests were used for laboratory confirmation.

[2]Includes persons vaccinated with CanSino at any time during cohort follow-up from March–December 2021.

**Supplementary Table 2: Underlying medical conditions reported among participants with COVID-19 associated hospitalization or death -- Mexico, 2021**

| Underlying medical condition | Outcome (n, [%]) | |
|---|---|---|
| | Hospitalization (n=25) | Death (n=6) |
| Any underlying medical condition | 18 (72%) | 6 (100%) |
| Diabetes | 6 (24%) | 5 (83%) |
| Hypertension | 6 (24%) | 4 (67%) |
| Obesity | 6 (24%) | 3 (50%) |
| Pregnancy | 4 (16%) | 1 (17%) |
| Any renal disease | 3 (12%) | 0 |
| Cancer | 2 (8%) | 0 |
| Any lung disease | 1 (4 %) | 0 |
| Any cerebrovascular disease | 1 (4%) | 0 |
| Immunosuppression | 1 (4%) | 0 |

**Supplementary Table 3: Participants with laboratory-confirmed COVID-19, by type of laboratory test -- Mexico, 2021**

| Characteristic | Total positive | % | Type of laboratory confirmation | | | | |
|---|---|---|---|---|---|---|---|
| | | | Rapid antigen | % | RT-PCR | % | p-value[1] |
| **Total** | 2250 | 100% | 2104 | 100% | 146 | 100% | |
| **Vaccination status** | | | | | | | |
| Vaccinated with CanSino | 1855 | 82% | 1742 | 83% | 113 | 77% | 0.09 |
| Unvaccinated | 395 | 18% | 362 | 17% | 33 | 23% | |
| **Sex** | | | | | | | |
| Female | 2142 | 95% | 2001 | 95% | 141 | 97% | 0.42 |
| Male | 108 | 5% | 103 | 5% | 5 | 3% | |
| **Median age (IQR)** | | 33 (27 – 40) | | 33 (27 – 40) | | 31 (27 – 39) | 0.29 |
| **Age group** | | | | | | | |
| 18-49 | 2104 | 94% | 1952 | 93% | 137 | 94% | 0.63 |
| 50+ | 146 | 6% | 152 | 7% | 9 | 6% | |
| **State** | | | | | | | |
| Distrito Federal | 343 | 15% | 324 | 15% | 19 | 13% | <0.001 |
| Aguascalientes | 31 | 1% | 31 | 1% | 0 | 0% | |
| Baja California | 56 | 2% | 48 | 2% | 8 | 5% | |
| Baja California Sur | 59 | 3% | 55 | 3% | 4 | 3% | |
| Campeche | 5 | 0% | 3 | 0% | 2 | 1% | |
| Chiapas | 26 | 1% | 19 | 1% | 7 | 5% | |
| Chihuahua | 46 | 2% | 45 | 2% | 1 | 1% | |
| Coahuila | 72 | 3% | 70 | 3% | 2 | 1% | |
| Colima | 39 | 2% | 39 | 2% | 0 | 0% | |
| Durango | 24 | 1% | 24 | 1% | 0 | 0% | |
| Estado De México | 74 | 3% | 69 | 3% | 5 | 3% | |
| Guanajuato | 97 | 4% | 92 | 4% | 5 | 3% | |
| Guerrero | 25 | 1% | 25 | 1% | 0 | 0% | |
| Hidalgo | 29 | 1% | 29 | 1% | 0 | 0% | |
| Jalisco | 112 | 5% | 104 | 5% | 8 | 5% | |
| Michoacán | 109 | 5% | 106 | 5% | 3 | 2% | |
| Morelos | 35 | 2% | 35 | 2% | 0 | 0% | |
| Nayarit | 83 | 4% | 83 | 4% | 0 | 0% | |
| Nuevo León | 150 | 7% | 138 | 7% | 12 | 8% | |
| Oaxaca | 35 | 2% | 34 | 2% | 1 | 1% | |
| Puebla | 31 | 1% | 24 | 1% | 7 | 5% | |
| Querétaro | 42 | 2% | 39 | 2% | 3 | 2% | |
| Quintana Roo | 47 | 2% | 45 | 2% | 2 | 1% | |
| San Luis Potosí | 50 | 2% | 45 | 2% | 5 | 3% | |
| Sinaloa | 137 | 6% | 122 | 6% | 15 | 10% | |
| Sonora | 97 | 4% | 92 | 4% | 5 | 3% | |
| Tabasco | 22 | 1% | 21 | 1% | 1 | 1% | |
| Tamaulipas | 108 | 5% | 100 | 5% | 8 | 5% | |

| | | | | | | |
|---|---|---|---|---|---|---|
| Tlaxcala | 4 | 0% | 4 | 0% | 0 | 0% |
| Veracruz | 143 | 6% | 134 | 6% | 9 | 6% |
| Yucatán | 92 | 4% | 82 | 4% | 10 | 7% |
| Zacatecas | 27 | 1% | 23 | 1% | 4 | 3% |

Abbreviations: RT-PCR, reverse transcription polymerase chain reaction

[1]P-value by chi-squared test (categorical) or Mann-Whitney test (continuous).

**Supplementary Table 4: Effectiveness of CanSino vaccine in preventing laboratory-confirmed COVID-19, hospitalization, by type of laboratory test -- Mexico, 2021**

| Test type | SARS-CoV-2 infections | VE estimate (95% CI) | | Hospitalizations | VE estimate (95% CI) | | Deaths |
|---|---|---|---|---|---|---|---|
| | N | Unadjusted | Adjusted[1] | N | Unadjusted | Adjusted[1] | N |
| Any test | 2250 | 14% (3-23%) | 20% (10-29%) | 25 | 73% (36-88%) | 76% (42-90%) | 6 |
| Rapid antigen test | 2104 | 20% (10-29%) | 30% (20-38%) | 15 | 81% (42-94%) | 91% (64-98%) | 6 |
| RT-PCR | 146 | 16% (-27-44%) | 16% (-36-48%) | 10 | 51% (-90-88%) | 75% (-21-95%) | 0 |

Abbreviations: RT-PCR, reverse transcription polymerase chain reaction; VE, vaccine effectiveness

[1]Adjusted for age, sex, state, and local viral circulation.

Stratified VE estimates for death are not presented as no deaths occurred among persons with RT-PCR-confirmed illness.
